# Disruption of maternal IgA by prenatal antibiotics precedes intestinal *E. coli* colonization and late-onset sepsis in neonates

**DOI:** 10.1101/2023.09.04.23295031

**Authors:** Katya McDonald, Danielle Browning, Kara G. Greenfield, Kathryn Lin, Josephine Meier, Matthew Butoryak, Alexandria Sasaki, Eva Fandozzi, Tim Hand, Christina Megli, Kathryn A. Knoop

## Abstract

Neonates, particularly prematurely born neonates, are particularly vulnerable to enteric pathogens. Antibiotics are widely administered during pregnancy for off-label use and prevention, but the effect on neonatal physiology has not been well characterized. Maternally derived IgA provided in milk is a predominant protective measure in the intestinal lumen of the nursing offspring. The connection between the maternal intestine and mammary IgA responses has been observed, but understanding how clinical use of antibiotics effects mammary IgA remains unclear. Here we find that prenatal cephalosporin antibiotics, but not penicillins, decrease mammary IgA in dams, and limit the availability of IgA within the lumen of nursing offspring. Decreased IgA correlated with an outgrowth of commensal *E. coli* and increased colonization and translocation of pathogen *E. coli.* Moreover, antenatal cephalosporin administration was associated with increased mortality in a model of LOS that was associated with decreased mammary IgA. Futhermore, in a clinical cohort of at-risk infants, maternal broad spectrum antibiotic treatment, including cephalosporins, increased the risk of LOS in neonates in comparison to penicillin-based treatments. This was associated with reduced IgA in the milk, and we propose maternal administration of select antibiotics could disrupt mammary IgA, leading to increased risk of LOS in infants by allowing for pathogen colonization in the neonatal intestine.

## Introduction

Late-onset neonatal sepsis (LOS) resulting from bloodstream infections occurring 7 days following delivery, are a major contributor to neonatal mortality. Very low birth weight (VLBW, <1500 grams) infants are particularly at risk for LOS, with an incidence of 10%, and a mortality rate of 18% (1). In a substantial portion of LOS, the causative bacterial pathogen can be found residing in the neonatal intestine as part of the intestinal microbial community. This implies intestinal translocation as the bacterial source (2–6). Accordingly, multiple animal models of LOS show gut-administered enteric pathogens can breech the intestinal barrier and secondarily disseminate causing systemic disease (7–9). Thus, the route of LOS is in stark contrast to the route typically associated with early-onset sepsis. Early-onset neonatal sepsis, occurring >72 hours following delivery, is thought to be the result of a bacterial infection introduced *in utero* or during the delivery process. These distinctions indicate there is a clear need to understand the causative risks for LOS compared to early-onset neonatal sepsis.

Human studies have also demonstrated that enteral route of infection contributes to morbidity and mortality (10) of LOS. *Escherichia coli* is the second most common causative pathogen of LOS. Similar to what is seen in animal models, in human neonates *E. coli* has been found to originate from the neonatal intestine in a substantial portion of LOS-causative bloodstream infections (11–14). As *E. coli* is a common member of the neonatal intestinal microbiota, luminal protection within the neonatal intestine is key to preventing opportunistic pathogens from disseminating systemically.

Maternal milk contains numerous factors that are protective against LOS, including oligosaccharides, secretory antibodies, growth factors and lactoferrin that contribute to protection against infection in the neonate (15–17). Overall, maternal milk has been linked with lower infant death, hospitalization risk and risk of sepsis (18–21). Immunoglobulin A (IgA) has long been known to be a valuable component of maternal milk in providing protection to the neonate by binding pathogens (22, 23). Maternal IgA cross-reacts to a variety of gut resident pathogens, including viruses, bacteria, and parasites (24–26), and shapes the developing neonatal microbiome (27–29). Maternal IgA originates from plasma cells that migrate from the maternal gastrointestinal tract to the mammary glands (30, 31), delineating the “Gut-Mammary Axis” (32). Recent observations in mice has shown maternal antibiotics throughout lactation can decrease mammary IgA through influence of the gut microbiota (31), yet how this affects the nursing offspring remains unclear.

While neonatal antibiotic use has been shown to increase risk to LOS, perinatal maternal antibiotics can also affect the neonate (33, 34). Antibiotics are prescribed in 25-30% of all pregnancies, making them the most common medication given to pregnant women (35). Indeed, in contemporary management of Preterm Premature Rupture of Membranes (PPROM), it is recommended that all pregnant patients receive antibiotics to prolong pregnancy in the absence of infection. Previous studies have demonstrated that broad spectrum antibiotic administration for patients who present with PPROM were associated with increased LOS, NEC and adverse neonatal outcomes (36). Despite this, there are clinical trends towards use of additional broad-spectrum antibiotics without prospective safety studies, culture guided treatments and without neonatal follow up. Use of broad-spectrum antibiotics is clinically relevant given the increasing rates of perceived penicillin allergy (37). When the recommended penicillin-based regimens are unavailable due to an allergy, selection of the antibiotics is left up to the clinician, yet not all antibiotic regimens have been studied for efficacy, especially when considering neonatal outcomes.

In this study, we sought to determine if disruption of mammary IgA from maternal antibiotic administration would increase risk of enteric pathogens and LOS. Broad spectrum antibiotics, specifically cephalosporins, lead to significantly reduced maternal IgA passed to the nursing neonate with decreased binding of *E. coli* in the nursing offspring, increasing susceptibility of LOS in mice. Clinically, we observed a strong association with alternate, broader spectrum maternal antibiotic use and the development of LOS in a high-risk cohort of premature infants who were exposed to maternal breast milk. Thus, maternal antibiotics may increase risk of LOS in infants through the reduction of pathogen-reactive IgA in maternal milk.

## Results

### Maternal Cephalosporin is associated with Reduced Mammary IgA

Previous studies observed that antibiotic depletion of the maternal microbiota during pregnancy and throughout lactation resulted in significantly decreased mammary IgA (31). To investigate the impact of perinatal antibiotics on neonates, we administered antibiotics during pregnancy. A variety of clinical-relevant dosages of antibiotics (including amoxicillin, ampicillin, cephalexin, metronidazole, and ceftazidime) were administered to pregnant dams via the drinking water for seven days: from embryonic day 14 until delivery. Mice were then placed on regular drinking water following delivery. On postnatal day 2 (PN2), IgA was significantly reduced in mammary glands following cephalexin and ceftazidime treatment, but not other penicillin-based antibiotics or metronidazole (Fig. 1A). This reduction was specific to the mammary glands, and neither cephalexin nor ceftazidime significantly decreased IgA in the serum (Fig. 1B). In the pups born to cephalexin or ceftazidime-treated dams, there was a significant decrease in intestinal IgA (Fig. 1C) but not serum IgA (Fig. 1D). We next assessed pups at PN7, and found decreased IgA in both the mammary glands and neonatal intestinal tracts following prenatal cephalexin administration (Fig. 1E,F). This difference was pronounced throughout the gastrointestinal tract in the pups, with significantly less IgA in the stomach, small intestine, and stool, without any significant difference in serum IgA (Fig. 1F). In contrast, there was no reduction of IgG in the dam or pups (Fig. 1G,H). Thus, maternal use of cephalexin and ceftazidime reduces the IgA in both the mammary glands and provided to the nursing pups, without limiting systemic IgA or IgG.

**Figure 1:**
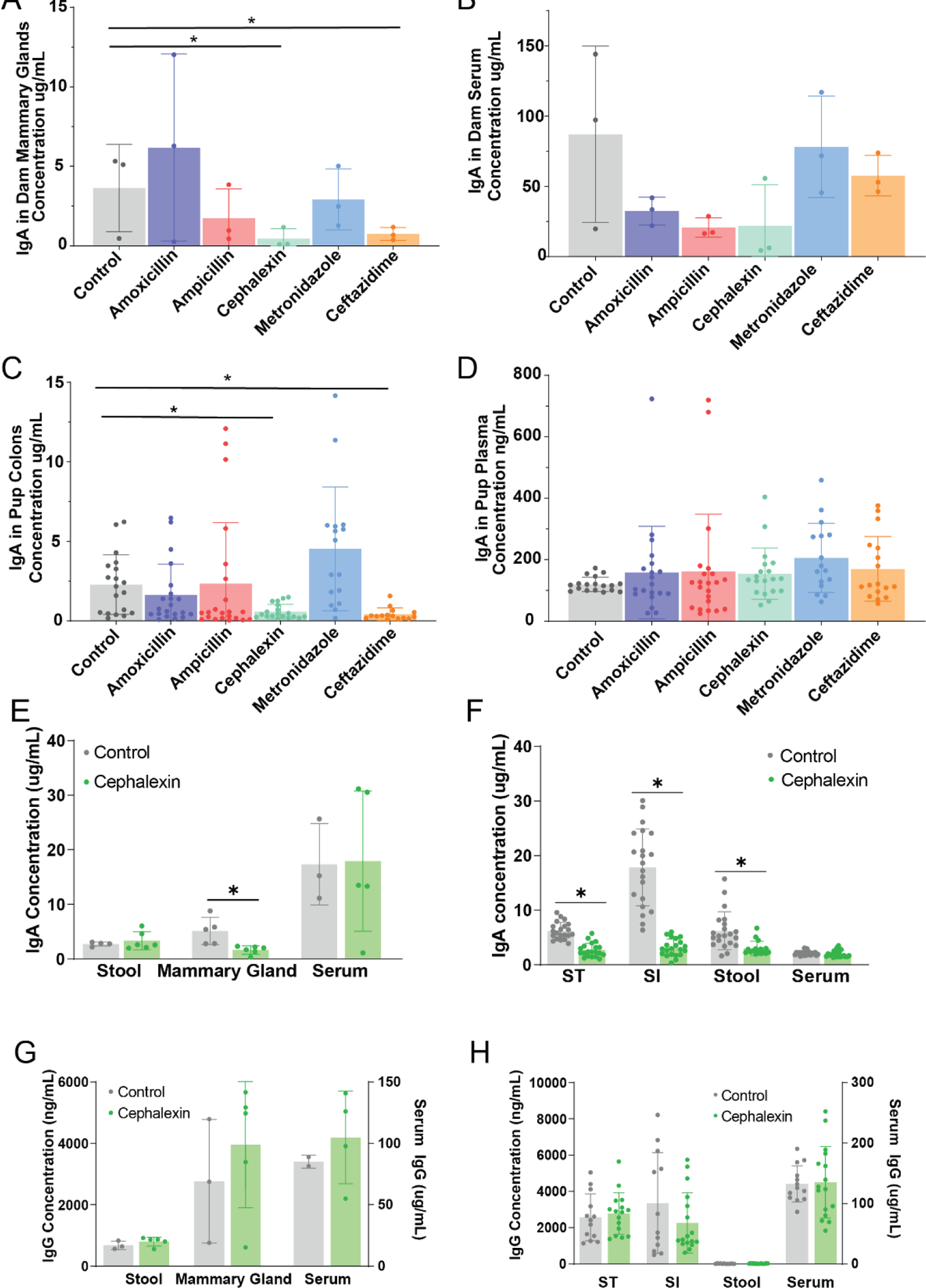
Cephalosporins decrease mammary IgA and luminal IgA in nursing pups. Stated antibiotics were administered to pregnant dams in drinking water from ED14 to delivery of litter, and tissue was analyzed on PND2. Total IgA was measured by ELISA from A) mammary glands of dams, B) serum of dams, C) colon contents of pups, and D) plasma of pups. Cephalexin was administered to pregnant dams in drinking water from ED14 to delivery of litter, and tissue was analyzed on PND7. Total IgA was measured by ELISA from E) stool, mammary and serum of dams, and F) stomach contents (ST), small intestinal contents (SI), stool, and serum of pups. Total IgG was measured by ELISA from G) stool, mammary and serum of dams, and H) stomach contents (ST), small intestinal contents (SI), stool, and serum of pups. One way ANOVA was used to calculate statistical significance. N=3 dams and corresponding litters per group.

### Reduced Mammary IgA is associated with Disruptions of the Maternal Microbiota

It has been reported that the concentration of IgA in murine milk differs between murine strains, potentially due to the difference in polyreactive IgA across strains, with BALB/c mice having increased IgA compared to C57BL/6 mice (38, 39). We did not observe a decrease in mammary IgA in BALB/c dams following cephalexin treatment (Fig. 2A). Similarly, BALB/c pups from cephalexin-treated dams did not have decreased intestinal IgA (Fig. 2B). We therefore used the distinct responses of C57BL/6 and BALB/c dams to cephalexin to study the effects of decreased mammary IgA, while controlling for off-target effects of cephalexin.

**Figure 2:**
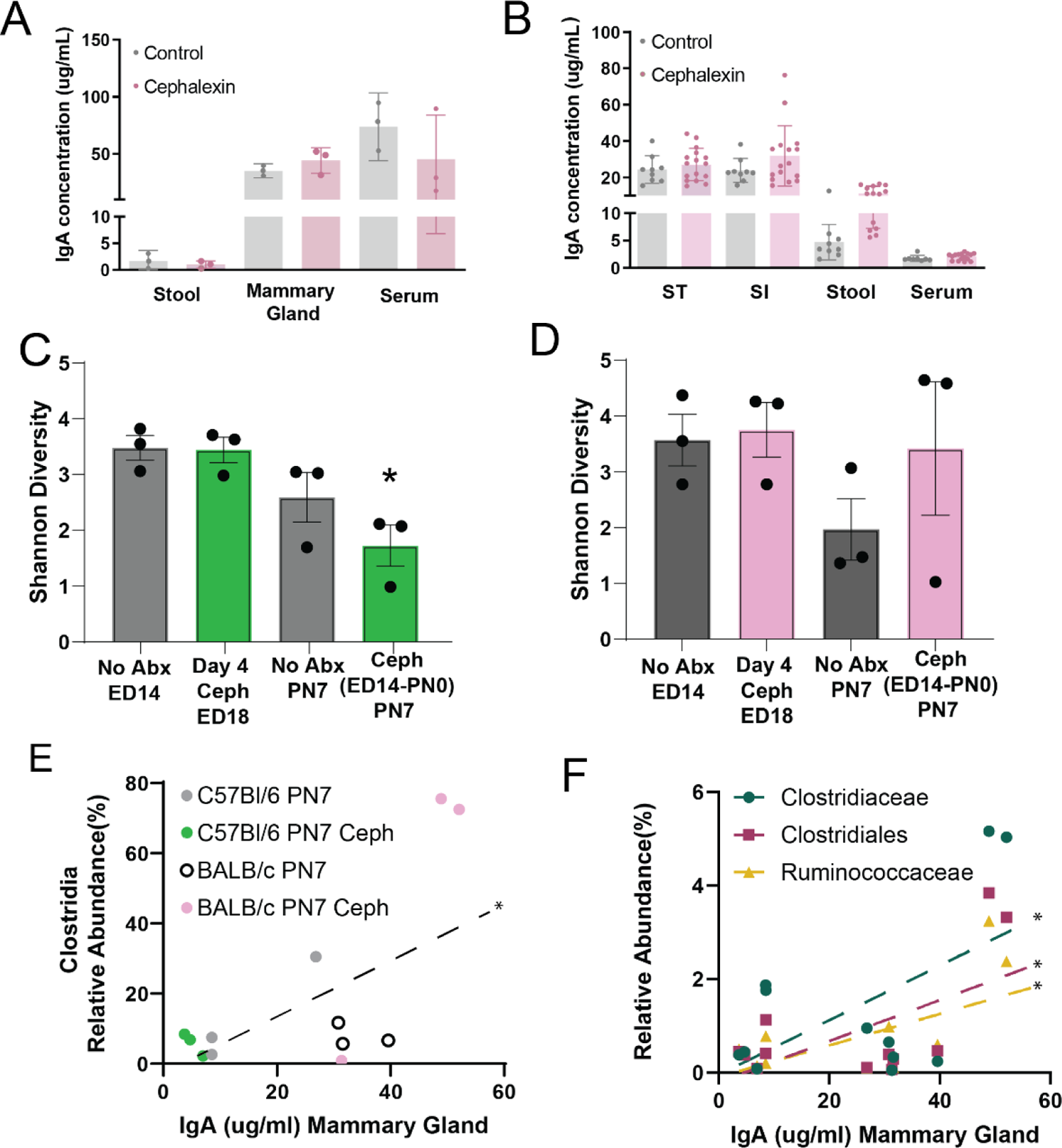
Cephalexin-induced decrease in mammary IgA is associated with decreased maternal microbial diversity. Cephalexin was administered to pregnant dams in drinking water from ED14 to delivery of litter, and tissue was analyzed on PND7. Total IgA was measured by ELISA from A) stool, mammary and serum of dams and B) stomach contents (ST), small intestinal contents (SI), stool, and serum of pups. C,D) Shannon diversity of 16S analysis of stool from C) C57BL/6 dams and D) BALB/c dams before, during and following cephalexin treatment. E) Relative abundance of clostridia class taxa correlated with IgA in mammary gland on PN7. Pearson’s correlation of all Clostrida relative abundance and all IgA p= 0.0102. F) Relative abundance of clostridia family taxa correlated with IgA in mammary gland on PN7. Pearson’s correlation for all IgA and: Clostridiaceae p=0.0500, Clostridiales p=0.0286, Ruminococcaceae p=0.0226. One-way ANOVA (A-D) and Pearson’s correlation (E,F) was used to calculate statistical significance. N=3 dams per group.

To understand if microbial disruptions in the intestines of dams contribute to reduced mammary IgA, we assessed the microbiota of stool during and after antibiotic administration. Cephalexin administration began at ED14, and four days into cephalexin administration (ED18), *Lactobacilli* species were significantly reduced in both C57BL/6 and BALB/c mice (Supplementary Figure 1a,b). Interestingly, these species were largely restored by PN7 suggesting cephalexin treatment effects on the microbiota were transient (Supplementary Figure 1a,b). Beyond decreased *Lactobacilli* strains we observed the C57B/6 dams and BALB/c dams had different responses to the cephalexin treatment, appreciable at the class taxa level (Supplementary Figure 1c). There was an expansion of the *Bacteroidia*, particularly *Muribaculaceae* in the C57BL/6, while there was an expansion of the *Clostridia*, particularly *Lachnospiraceae*, in the BALB/c (Supplementary Figure 1c). Ultimately, these differences resulted in reduced Shannon diversity in the lactating C57Bl/6 dams following cephalexin (Fig. 2C), but not the BALB/c dams (Fig. 2D). Since we observed differences in the microbial trajectories between BALB/c and C57BL/6 dams following cephalexin, we asked if mammary IgA correlated with specific taxa. We found a positive correlation of mammary IgA with *Clostridia* species, including *Clostridiaceae, Clostridiales,* and *Ruminococcaceae* (Fig. 2E,F). Taken together, these data suggest microbial disruptions, including decreased diversity, to the C57B/6 dams resulting from cephalexin is associated with decreased mammary IgA.

### Reduced mammary IgA is associated with increased E. coli colonization in mice

To understand the effect of decreased mammary IgA on nursing offspring, we assessed the microbiota of pups on PN7. Though the cephalexin administered to the dams was exclusive to pregnancy, we observed a noticeable shift in the microbiota of the C57BL/6 pups. Principal component analysis comparing the microbiota of the pups found the cephalexin treatment had distinct effects on BALB/c pups as compared to C57BL/6 pups (Supplementary Figure 2a). Pups of cephalexin-treated dams had the most observable microbial difference: an increase in *Enterobactericeae* (Supplementary Figure 2b) and a significant increase in the relative abundance of *E. coli* (Fig. 3A), suggesting the reduction in maternal IgA was associated with an increase in *E. coli* in the offspring. Notably, in BALB/c pups, Gammaproteobacteria, and *E. coli* was not a major component of the microbiota either with or without cephalexin (Fig. 3A). Thus, the increased *E. coli* in C57BL/6 pups of cephalexin treated dams is associated with decreased mammary IgA, and unlikely to be due to other off-target effects of cephalexin. IgA from the stomach contents of PN7 pups was capable of binding pathogenic *E. coli*, and this was decreased in PN7 pups from cephalexin-treated dams (Fig. 3B). Interestingly, cross-reactivity of another bacterial pathogen, group B streptococcus (GBS), remained unchanged after cephalexin treatment, suggesting a selective decrease in mammary IgA able to bind *E. coli*. The relative abundance of *E. coli* found in the pup stool negatively correlated with the amount of IgA in their small intestine (Fig. 3C). Thus, decreased maternal IgA resulted in expansion of *E. coli* in the nursing offspring.

**Figure 3:**
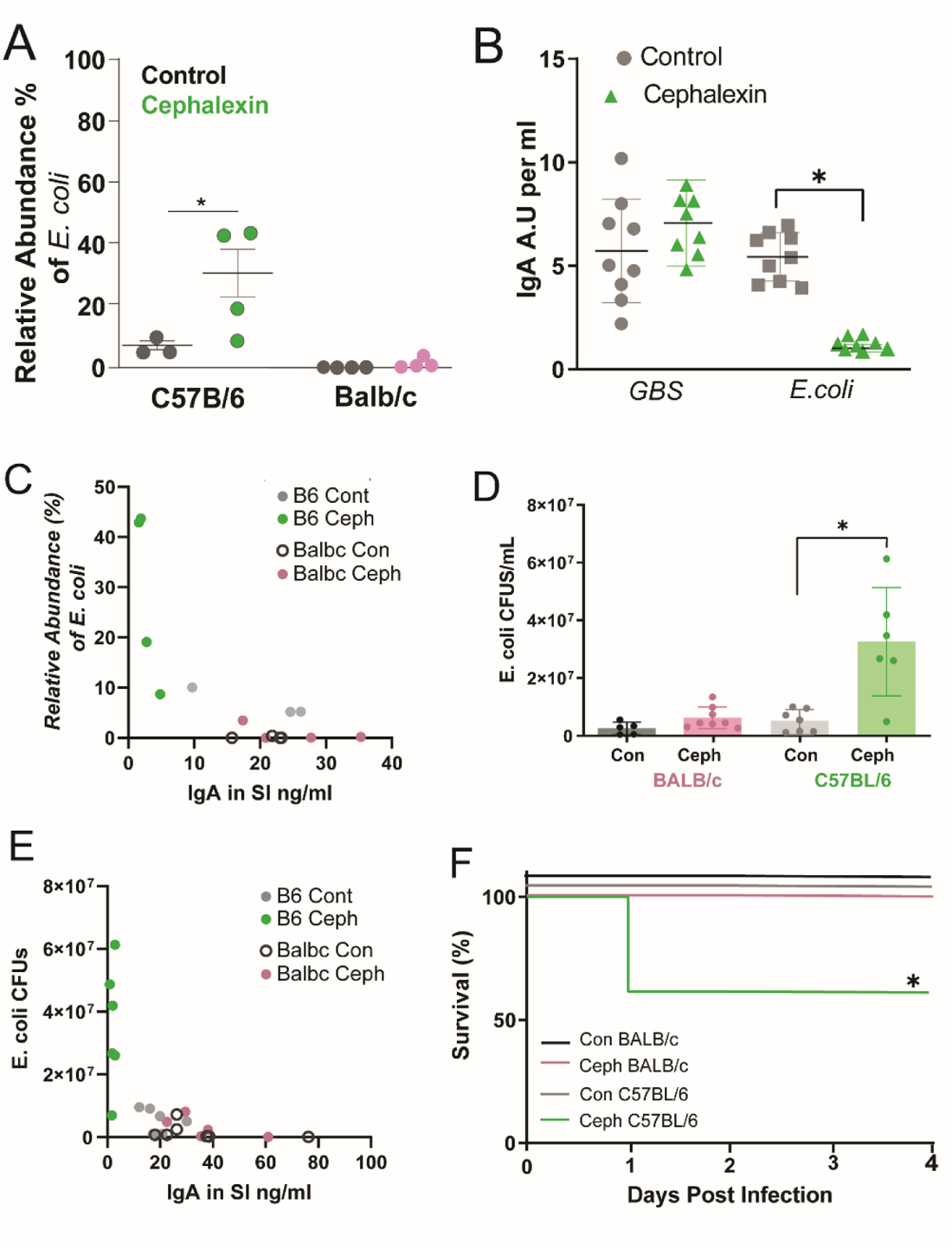
Cephalexin-induced decrease in mammary IgA is associated with increased E. coli colonization and LOS. A) Relative abundance of *E. coli* based on 16S analysis on stool of pups. B) Cross-reactive IgA from the stomach contents of pups following dam cephalexin treatment was measured against group B Streptococcus (GBS) and *E. coli*. C) Relative abundance of *E. coli* in the stool of neonates correlated with the concentration of IgA in the small intestine of corresponding samples. D-E) 10^6 CFUs of pathogenic BSI-B *E. coli* were gavaged to pups. D) CFUs from stool were plated 48 hours following infection. E) CFUs of *E. coli* in the stool of neonates correlated with the concentration of IgA in the small intestine of corresponding samples. F) Survival of pups following BSI-B gavage on PN5. One way ANOVA was used to calculate statistical significance, A,B,D,E, G. Kaplan Meier Curve was used in F. N=3 dams and corresponding litters per group.

### Reduced mammary IgA is associated with increased E. coli LOS in mice

We next asked if maternal IgA would limit pathogenic *E. coli* in pups following direct infection. To do this, we used a pathogenic ST131 *E.coli* isolated from the bloodstream of a neonatal LOS patient, bloodstream isolate B (BSI-B) (17). This strain of *E. coli* has been shown to translocate from the intestine resulting in bacteremia in a model of neonatal sepsis (17). PN5 pups from dams treated with or without cephalexin were orally inoculated with 10^6^ CFUs of BSI-B. 48 hours following oral infection, there was a significant increase in CFUs of *E. coli* in the stool of pups from cephalexin-treated C57BL/6 dams compared to pups from untreated C57BL/6 dams or those from BALB/c dams (Fig. 3D). As in uninfected mice, the colonization of *E. coli* in the pup stool negatively correlated with the amount of IgA in the small intestine (Fig. 3E). Finally, we observed increased mortality in the pups from cephalexin-treated C57BL/6 dams (40%) compared to both untreated dams and cephalexin treated BALB/c dams (Fig. 3F). Thus, reduced mammary IgA is associated with increased pathogenic *E. coli* colonization in the intestine and mortality in a model of LOS.

### Antenatal cephalosporin and non-beta lactam use is associated with increased rates of LOS in humans

To determine if there was a clinical association of antenatal antibiotic exposure and adverse neonatal outcomes, we generated a cohort of neonates born to mothers with PPROM (Fig. 4). The recommended regimen following PPROM is ampicillin IV for 48 hours followed by amoxicillin orally for 7 days in combination with a macrolide antibiotic (traditionally erythromycin, and not azithromycin). We identified two groups of patients: those receiving the traditional regimen of antibiotics (ampicillin/amoxicillin) and those receiving non-traditional antibiotics, through ICD9/ICD10 codes in the maternal chart and detailed maternal and neonatal chart review (Fig. 4a). Non-traditional antibiotics including either non-beta lactams or cephalosporins, both broader spectrum antibiotic regimens. There was no difference in pregnancy latency, gestational age at delivery, gestational age at PPROM, number of days that patient received antibiotics or breast milk feeding of neonates (Table 1). When assessing neonatal outcomes, there was no difference in select neonatal outcomes including early onset sepsis and NEC>2A. However, there was a significant increased risk of late onset sepsis with exposure to non-traditional antibiotics with an OR of 4.836 (p<0.0008) (Table 1).

**Figure 4:**
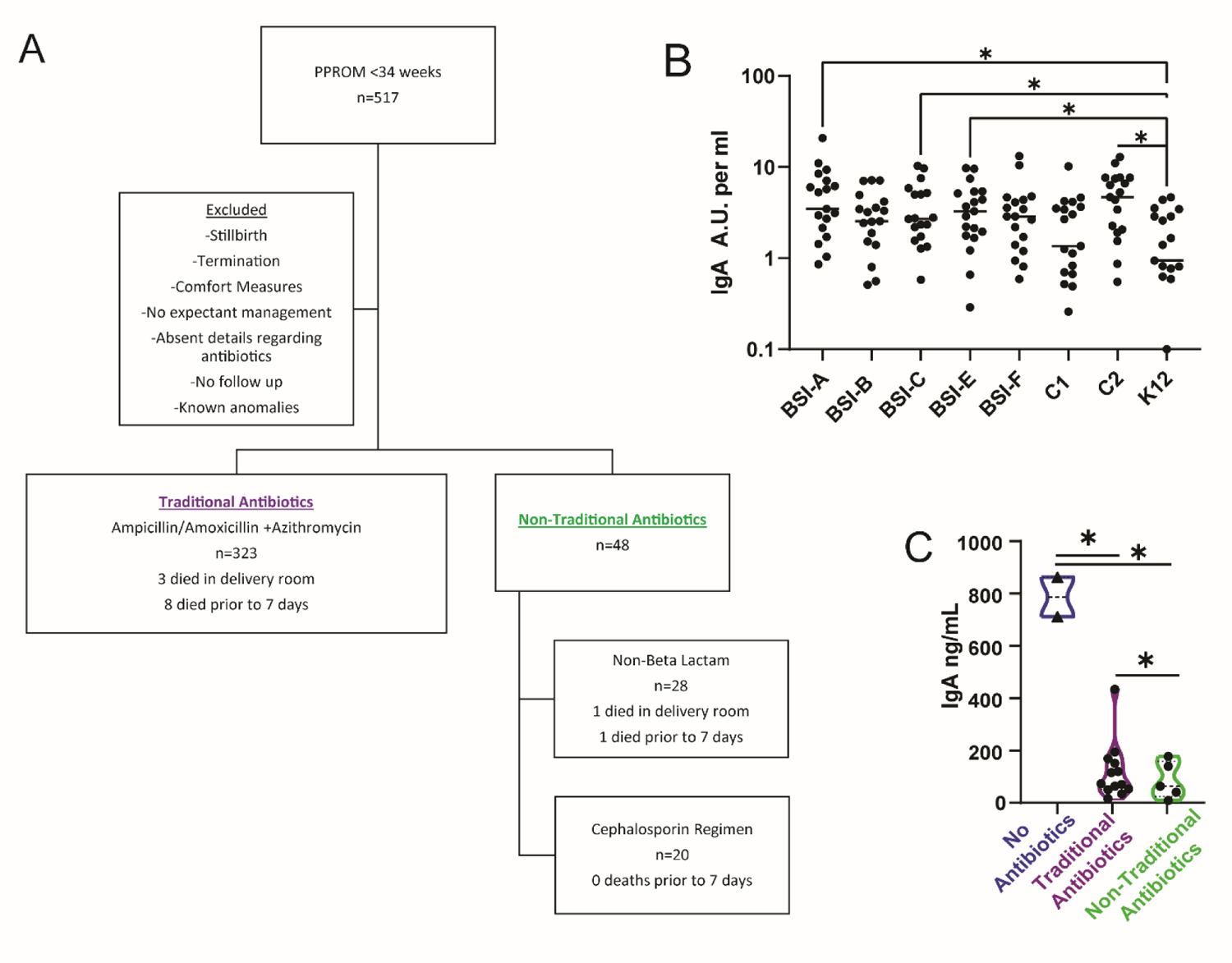
Non-traditional antibiotics is associated with decreased mammary IgA. A) Design of clinical cohort for the study of maternal antibiotics following PPROM (Table 1,2). B) Human milk cross reacts to *E. coli* commensals and pathogens. Cross-reactive IgA from human donor milk was quantified against indicated *E. coli* isolates. N=20 donor milk specimens. C) Total IgA in human milk following maternal antibiotics following PPROM. One-way ANOVA was used to calculate the statistical significance (B, C).

**Table 1:** Neonatal Outcomes following Maternal Antibiotics and PPROM,*Death, LOS, NEC, BPD/CLD, grade 4-3 IVH.

|  | Traditional Antibiotics<br>n=320 | Non-traditional Antibiotics<br>n=49 | OR (CI), p value |
| --- | --- | --- | --- |
| GA at PPRM | 208.3 | 206 | p=0.55 |
| GA at delivery | 217.0 | 215.4 | p=0.65 |
| No breast milk exposure | 19 (6.1) | 1 (2.2) | p=0.48 |
| Neonatal death | 13 (4.0) | 4 (8.3) | 2.168 (0.7422-6.902), p=0.18 |
| EOS | 15 (4.8) | 1 (2.1) | .4348 (0.040-2.721),p=0.41 |
| LOS | 11 (3.5) | 7 (14.9) | **4.836 (1.856-13.39), p=0.0008** |
| NEC≥2A | 17 (5.4) | 4 (8.5) | 1.631 (.5716-4.715), p=0.39 |
| LOS/NEC | 27 (8.6) | 9 (19.2) | **2.526 (1.057-5.599), p=0.02** |
| Composite outcome * | 145 (44.9) | 25 (52.1) | 1.334 (0.7458-2.415) p=0.35 |
| Table 1: Neonatal Outcomes following Maternal Antibiotics and PPRM,*Death, LOS, NEC, BPD/CLD, grade 4-3 IVH |  |  |  |

A sub-analysis of the non-traditional antibiotic regimen demonstrated that there was no difference between the cephalosporin regimen and non-beta lactam regimens with similar rates of late onset sepsis in both cohorts (Table 2). Positive blood cultures were also more likely in the non-traditional regimen in the work-up for late onset sepsis (6.38 vs 1.27%, p<0.05). Composite neonatal morbidity was higher in the non-traditional antibiotic regimens, but this did not achieve statistical significance. This data demonstrates that non-traditional antibiotic regimens, including cephalosporin-based regimens, are associated with increased rates of LOS.

**Table 2:** Neonatal outcomes following Maternal Antibiotics and PPROM,*Death, LOS, NEC, BPD/CLD, grade 4-3 IVH.

|  | Traditional<br>n=319 | Cephalosporin<br>n=20 | Non-beta lactam<br>n=28 |
| --- | --- | --- | --- |
| <u>GA at PPROM</u> | 208.3 | 206.5 | 205.6 |
| <u>Latency days</u> | 8.7 | 11.9 | 7.8 |
| <u>GA at delivery</u> | 217.0 | 218.4 | 231.5 |
| <u>Neonatal death</u> | 13 (4) | 1 (5) | 3 (6.3) |
| <u>Composite Neonatal Morbidity</u> | 145 (44.9) | 11(55.0) | 14(50.0) |
| <u>EOS</u> | 15 (4.7) | 0 | 1 |
| <u>LOS/SIP</u><br>Culture positive | 10 (3.2)<br>4 | 3 (15)<br>1 | 4 (14.8)<br>2 |
| <u>NEC ($\geq 2A$)</u> | 17 (5.1) | 1 (5) | 3 (11.1) |
| <u>NEC<2A</u> | 6 | 2 | 0 |
| <u>LOS/NEC $>2A$</u> | 27 (8.6) | 4 (20) | 5 (18.5) |
| <u>Other postnatal infection</u> | 15 (4.7) | 1 (5.0) | 2 (7.4) |
| Pneumonia/Respiratory | 4 | 0 | 0 |
| Diaper or Oral Candidiasis | 3 | 1 | 0 |
| UTI | 6 | 0 | 1 |
| Omphalitis/ cellulitis | 2 | 0 | 1 |

A subanalysis demonstrated that 2 neonates who developed LOS in the traditional antibiotics cohort were fed donor breast milk, the remainder were fed with maternal breast milk that was fortified as needed. There was incomplete data for one neonate who was treated for LOS in the non-traditional cohort. In this context, a comparison was made between maternal breastmilk exposed infants in the traditional and nontraditional regimens. There remained a substantial increase in the cases of LOS in the nontraditional cohort (13.3% versus 3.1%, OR 4.789 p=0.0084).

We then looked at the ability for mammary IgA to bind to *E. coli*. Human IgA from donor human milk was capable of binding to a variety of *E. coli* strains (Fig. 5B), including both pathogenic and commensal *E. coli* from neonates. We observed significantly more binding of pathogenic *E. coli* when compared to the lab strain K12 *E. coli*, suggesting maternal IgA may have improved cross-reactivity to intestinal-derived *E. coli*.

We then looked at IgA from human milk in a cohort of mothers who were administered antenatal antibiotics and had preterm deliveries. We observed a significant difference by ANOVA in IgA in milk from mothers treated with non-traditional broad-spectrum antibiotics, or traditional antibiotics, as compared to no antibiotics. This suggests maternal IgA may be disrupted by maternal antibiotics; however, our cohort did not have enough cases to discern differences between individual antibiotics (Fig. 5C). Taken together, we propose maternal IgA can limit intestinal colonization of *E. coli* in the nursing offspring and decreased maternal IgA following broad spectrum prenatal antibiotics increases risk of neonates to LOS. Our data have applications in clinical management as well as in understanding the pathophysiology of late onset sepsis in neonates.

## Discussion

PPROM is responsible for approximately 1/3 of all preterm birth and is a substantial risk factor for complications associated with prematurity. The landmark Mercer trial on PPROM suggested antibiotic management prolongs gestation and neonatal outcomes (40). However, there was a lack of extensive neonatal follow up past discharge or transfer, and only early-onset sepsis was included in the primary outcome, defined as occurring <72 hours after delivery (40). This approach has been widely adopted in the United States and, in our cohort, patients universally received antibiotics and achieved a substantially longer mean latency than was reported in the initial trial. Alternative antibiotics to the initial regimen have not been thoroughly studied with the exception of the ORACLE trial that exchanged ampicillin for ampicillin/sulbactam and amoxicillin for amoxicillin/sulbactam (36). In this trial, the addition of beta-lactamases led to increased rates of adverse neonatal outcomes but additional studies on other antibiotic regimens are not well studied.

The use of non-traditional antibiotics in the setting of PPROM is often secondary to a perceived penicillin allergy. In a retrospective re-analysis of the MFMU BEAM trial, non-beta lactam antibiotics were associated with increased rates of maternal endometritis and increased rates of adverse neonatal outcomes but did not reach significance (41). Cephalosporins are currently recommended for use with PPROM in the setting of a low-risk penicillin allergy, and therefore our analysis is different in that we included cephalosporin use in the non-traditional antibiotic cohort and saw increased rates of LOS. When combined with the evidence in the ORACLE trial, our data suggests maternally provided broad-spectrum antibiotics including those in the beta-lactam class can have adverse effects on neonates beyond the initial 72 hours after delivery. The potential link between maternal antibiotic use and neonatal LOS is decreased mammary IgA in milk. Our data suggest more studies are needed when using maternally provided broader spectrum antibiotics in the absence of infection, particularly on neonatal outcomes, and the effects on human milk.

The connection between broad spectrum antibiotics, disrupted mammary IgA and neonatal intestinal health is further supported by our preclinical model: antenatal cephalosporin was associated with decreased maternal IgA in the mammary glands and gastrointestinal tract of the nursing neonates. Decreased maternal IgA has been associated with increased *Enterobacteriaceae* species that preceded necrotizing enterocolitis, another intestinally-related disease commonly seen in preterm infants, showing the potent ability of maternal IgA in limiting potential enteric pathogens in neonates (42). Here, we show that in addition to protection from enteric disease by limiting intestinal colonization, maternal IgA also protects against the systemic dissemination that results in sepsis. This protection was strongly associated with maternal IgA and mammary IgA cross-reacted to *E. coli* more than IgG, and we did not observe a decrease in maternal IgG in mammary glands or the lumen of pups following cephalexin treatment.

Maternal IgA was associated with limiting both commensal *E. coli* as part of the microbiota, and orally administered pathogenic *E. coli,* as both were expanded in C57BL/6 pups following cephalexin treatment. Prior observations have shown intestinal IgA can limit the growth of *E. coli*, and selective IgA deficiency in humans resulted in increased *E. coli* (43). Maternal IgA responses targeting *E. coli* have long been appreciated to be transferred to neonates in livestock mammals (22) and orally administered monoclonal IgA protect against enteric bacterial infections, including *E. coli* (44). *E. coli* colonization was not expanded in BALB/c mice, which had significantly more maternally-derived IgA compared to C57BL/6 pups, even following cephalexin treatment. There was no significant shift in *E. coli* within the intestines of dams, either between murine strains or after antibiotics. Thus, the expansion of *E. coli* in the pups is unlikely due to increased vertical transmission from the dams. We have previously shown that *E. coli* translocation from the intestine was significantly increased when maternally-supplied epidermal growth factor (EGF) and offspring-expressed EGF receptor was disrupted, allowing for the formation of goblet cell-associated antigen passages (9). This suggests multiple maternal components may work in tandem for the intestinal protection of neonate: IgA limiting pathogen colonization, and EGF limiting access through goblet cell-associated antigen passages.

The source of mammary derived IgA has been long connected to gut derived IgA responses in the mother. IgA-committed lymphoblasts from the mesenteric lymph node home to the mammary glands specifically during late pregnancy (30). These IgA responses can be induced in gut-associated lymphoid tissue, such as the Peyer’s patches, in response to specific bacteria, including *Parabacteroides goldsteinii, B. acidifaciens,* and *Prevotella buccalis* (31). Disruption of these bacterial species with vancomycin treatment during pregnancy and throughout lactation reduced mammary IgA (31); however, we did not observe a significant shift in those species in our model. We observed different microbial trajectories in the C57BL/6 mice compared to BALB/c mice in response to cephalexin and, ultimately, the decreased diversity in the microbiota and clostridial species in C57BL/6 mice correlated with decreased mammary IgA. Non-penicillin- and azithromycin-based regimens typically have broader activity against gram-negative enteric bacteria in obstetric patients (M. Shaffer and C. Megli, submitted) compared to penicillin-based regimens, but analysis into how traditional and non-traditional antibiotics given following PPROM affect the maternal intestinal microbiota is needed as antibiotics can have extensive effects on the microbiota.

The basis for the specificity in IgA capable of binding *E. coli* remains unclear. The decreased mammary IgA seemed to be directed to *E. coli*: we observed increased colonization of *E. coli*, and decreased binding of *E. coli*, following cephalexin treatment, and the decreased mammary IgA did not show a universal decrease in binding all bacteria as there was no decrease in cross-reactivity to GBS. While dams contained *E. coli* within the intestinal microbiota at low abundance, they were not immunized against *E. coli* to induce a specific response. Similarly, the human IgA from donor milk had the ability to cross-react to multiple *E. coli* species. Thus, naturally-induced IgA response can provide maternal protection. Naturally-induced IgA have low affinity but high cross-reactivity to a number of intestinal bacteria (45), and microbial exposure at the mucosal surfaces influence the B cell and IgA repertoire (46). Previous reports using auxotrophic *E. coli* suggest transient exposure to commensals can result in sustained IgA memory response in the absence of commensals (28). This IgA memory response can be disrupted following shifts to the microbiota (28). This has led to the speculation that IgA memory in response to commensals may be short-lived, with rapid turnover, and impaired expansion upon restimulation (47). In our model, cephalexin treatment, and the resulting antibiotic-induced shift, is occurring during gestation between ED14-ED21. This could coincide with a time in life when IgA producing plasma cells are expanding or trafficking to the mammary gland in preparation for lactation. Further exploration into how this exposure can inform mammary IgA responses, and how disruptions to the maternal intestinal microbiota can disrupt mammary IgA responses, should address this rapid crosstalk in the gut-mammary axis.

One limitation of this study is the small number of human milk samples from the clinical cohort of mothers following PPROM. While we observed a trend toward decreased IgA in maternal milk following antibiotics, more work is required to understand if there is a decrease in IgA following specific antibiotics, such as cephalosporins. Additionally, measuring IgA in colostrum and throughout lactation, tracking changes in the maternal microbiota, and assessing the binding reactivity of IgA following antibiotics will allow us to understand how maternal IgA may be affected by maternal antibiotic use. These data should be considered with the reports advocating for treatment of intrauterine infection using broad spectrum antibiotics and expectant management. Moreover, our data further suggest that the microbiome-immune axis is relevant to neonatal health. Further studies in how antibiotics can cause a perturbation of the microbiome-immune axis and lead to consequences in maternal-child health are warranted.

In conclusion, selective maternal antibiotic treatment perinatally can increase risk in neonates for LOS, as it reduces mammary IgA responsible for limiting pathogen colonization and expansion in the intestinal lumen of the nursing offspring. Our work builds on previous data showing the contribution of maternal IgA in influencing the developing microbiota (27) and how maternal antibiotic use can decrease mammary IgA (31) by identifying a functional outcome of the decreased mammary IgA in the nursing offspring. IgA binding of bacteria can create a positive feedback loop to promote future IgA responses (38, 39); so, disruption of this process may reduce future IgA responses against potential enteric pathogens in the offspring. Future directions should explore the gut-mammary axis to better understand how maternal antibiotics affect mammary IgA responses and neonatal health outcomes.

## Methods

### Patient inclusion

Patients were identified using ICD9/ICD10 codes for PPROM and delivering within the UPMC system between 2017-2020. Maternal and neonatal charts were reviewed, and multiple clinical variables and outcomes were collected and deidentified. Data were reviewed by a maternal fetal medicine specialist (CM) and a neonatologist (DB). Patients were excluded from analysis in the setting of previable delivery less than 23 weeks, stillbirth, incomplete information regarding antibiotic exposure and if they were not eligible for expectant management. Inclusion and exclusion criteria are demonstrated in Figure 4a. This study was approved through the University of Pittsburgh IRB board.

### Definition of clinical outcomes

Late onset sepsis was defined as diagnosis of sepsis greater than or equal to 7 days after delivery. Necrotizing enterocolitis was included if radiologic findings were consistent with necrotizing enterocolitis greater than 2A based on Bells Staging Criteria. Diagnosis of late onset sepsis and spontaneous intestinal perforation were included if patients were clinically diagnosed and treated. Neonates who underwent a diagnostic evaluation for late onset sepsis but did not complete treatment were not considered to have late onset sepsis.

### Mice

C57BL/6J and BALB/cJ mice were purchased from the Jackson Laboratory and bred in-house at the Mayo Clinic. All mice were stored in standard housing. All mouse husbandry and experiments involving mice were approved by the Mayo Clinic Institutional Animal Care and Use Committee (IACUC).

### Bacterial Isolates

*E. coli* strains BSI-A, BSI-B, BSI-C, BSI-D, BSI-E, and BSI-F are bloodstream isolates, along with non-pathogenic strains C1 and C2, which were isolated from stool from infants. (9) Commercial E. *coli* strain K12 (non-pathogenic) was purchased from the American Type Culture Collection (Manassas, VA, USA). All bacteria were cultured at 37°C in Lennox L (LB) Broth or Agar (Invitrogen, Waltham, MA, USA). Bacterial strains were made nalidixic acid resistant as previously described (9).

### E. Coli Infection

Mice were orally gavaged with 50 uL of PBS containing 10^6^ colony-forming units (CFUs) of BSI-B on day of life five. Mice were weighed at 24hrs post infection to assess the impacts of the infection. If mice had lost any weight at the 24hr check, they were euthanized and declared moribund. All surviving mice were euthanized at 48hrs post-infection.

### Antibiotic Treatments

14 days after mating (embryonic day 14), the male and female were separated. At this point, antibiotics were given to the female pregnant mouse via drinking water until delivery. This included amoxicillin (0.2mg/mL), ampicillin (0.4mg/mL), cephalexin (0.4mg/mL), metronidazole (0.4mg/mL) or ceftazidime (0.4mg/mL). Mice were then placed on regular drinking water following delivery of the litter.

### Quantification of Murine IgA/IgG or Human IgA

Murine IgA and IgG were quantified using the IgA Mouse Uncoated ELISA Kit and the IgG (Total) Mouse Uncoated ELISA Kit (Invitrogen). Samples were diluted in PBS, and assays were performed according to manufacturer’s instructions. The optical density was found using a BioTek 800 TS absorbance reader at 450nm. Concentrations of IgA and IgG were reported as ng/mL based on an intraassay standard curve. Human IgA was quantified using the Human IgA ELISA kit (Invitrogen). Concentrations of IgA were reported as ng/mL based on an intraassay standard curve.

### 16S Metagenomic Sequencing

Stool from pups and dams were taken from uninfected mice on PN7. Dam stool was sequenced per each individual dam, while stool from pups of the same litter were pooled for a representative litter sequence. Murine stool DNA was isolated using the PowerFecal Pro DNA Kit (QiAmp) and sequenced using the 16S Barcoding Kit 1-24 (SQK-16S024) on a GridION (Nanopore). Sequences were aligned using Centrifuge (v1.0.4) (48, 49). Data was analyzed using R (v4.2), and all mouse and human sequence data was removed during analysis. Data analysis included alpha diversity, principal component analysis, and taxonomical distribution.

### Tissue Collection

For DOL 5-7 pups, stool/colon contents were homogenized at 1mg/100µL PBS, stomach contents in 200µL PBS, and small intestine in 100µL PBS, and serum was collected via cheek puncture. For DOL 0-1 pups, colon, stomach, and small intestine homogenized in their entirety in 100µL PBS and serum was collected during euthanasia. For dam samples, stool was homogenized at 1mg/mL PBS, and mammary glands at approximately 0.2729 g/840µL PBS. The portion of mammary gland homogenized was slightly varied among samples, so all data is normalized by weight.

### Tissue CFUs

The spleen and mesenteric lymph node were homogenized in 300µL PBS, then serially diluted, and plated for CFUs on LB plates. For stool plating, 0.1 mg of stool was homogenized in 300µL PBS, then serially diluted, and plated for CFUs on LB plates. 20 ug/ml nalidixic acid was added to MacConkey agar for selective growth of nalidixic acid resistant BSI *E coli*.

### Cross Reactive IgA from Human Milk

Unpasteurized donor milk flagged for research use was obtained from the Minnesota Milk Bank for Babies. Total IgA was quantified using Uncoated ELISA Kit (Invitrogen). Assay was performed in triplicate. The optical density was found using a BioTek 800 TS absorbance reader at 450nm. Concentrations of IgA were reported as ng/mL based on an intraassay standard curve (provided by kit manufacturer). Cross-reactive IgA was measured using the IgA Human Uncoated ELISA Kit (Invitrogen). For the coating step, bacteria were grown up to 0.3 OD in LB Broth, spun down and resuspended in PBS, then heat-killed (via heat block, for at least one hour) and added to the plate (100µL a well). The rest of the experimental protocol followed the manufacturer’s instructions, data was reported as the absorbance units.

### Statistical Analysis

All statistical analysis was done using GraphPad Prism 6.0 (GraphPad Software Inc.). Data is reported as mean +/- standard deviation. Differences between two groups was analyzed using one-way ANOVAs, with Sidak’s posttest. For the clinical cohort, continuous variables were compared using students T test and dichotomous variables were compared using Chi-Squared Analysis.

## Author Contributions

KM, KG, KL, and JM performed antibiotic animal experiments, flow cytometry, ELISAs, and data analysis. CM oversaw clinical cohort design, data acquisition and analysis. EF performed the Human IgA ELISA. TH collected the maternal high risk milk samples DO and AS performed detailed chart review of clinical cohort. KK performed 16S analysis. CM and KK wrote initial manuscript, all authors discussed and edited the manuscript and contributed to figure preparation.

## Data Availability

All data produced in the present study are contained in the manuscript, except for metagenomic sequencing files, which are available upon reasonable request to the authors and will be publically available in the NIH GenBank SRA database.

Metagenomic 16S sequences will be deposited and available in the NIH GenBank SRA database.

## Notes

### Competing Interest Statement

The authors have declared no competing interest.

### Funding Statement

This study was funded by the NIH DK134366 (KK).

### Author Declarations

IRB of the University of Pittsburgh gave ethical approval for this work.

### Summary of Updates

Revised order of figures for clarity

