## Supplementary figures and images for "Disruption of maternal IgA by prenatal antibiotics precedes intestinal *E. coli* colonization and late-onset sepsis in neonates"

### Supplementary Figure 1

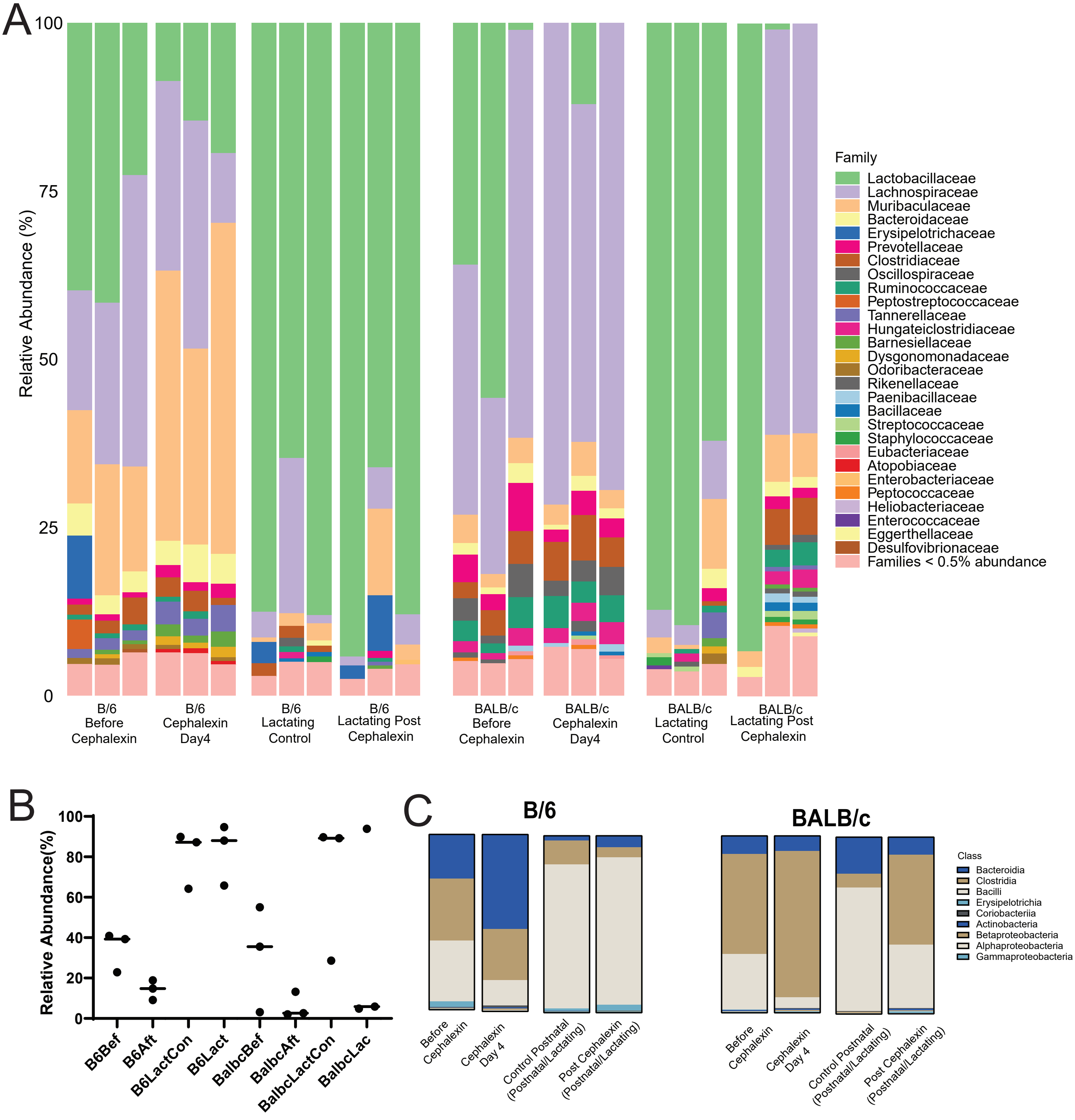

### Supplementary Figure 2

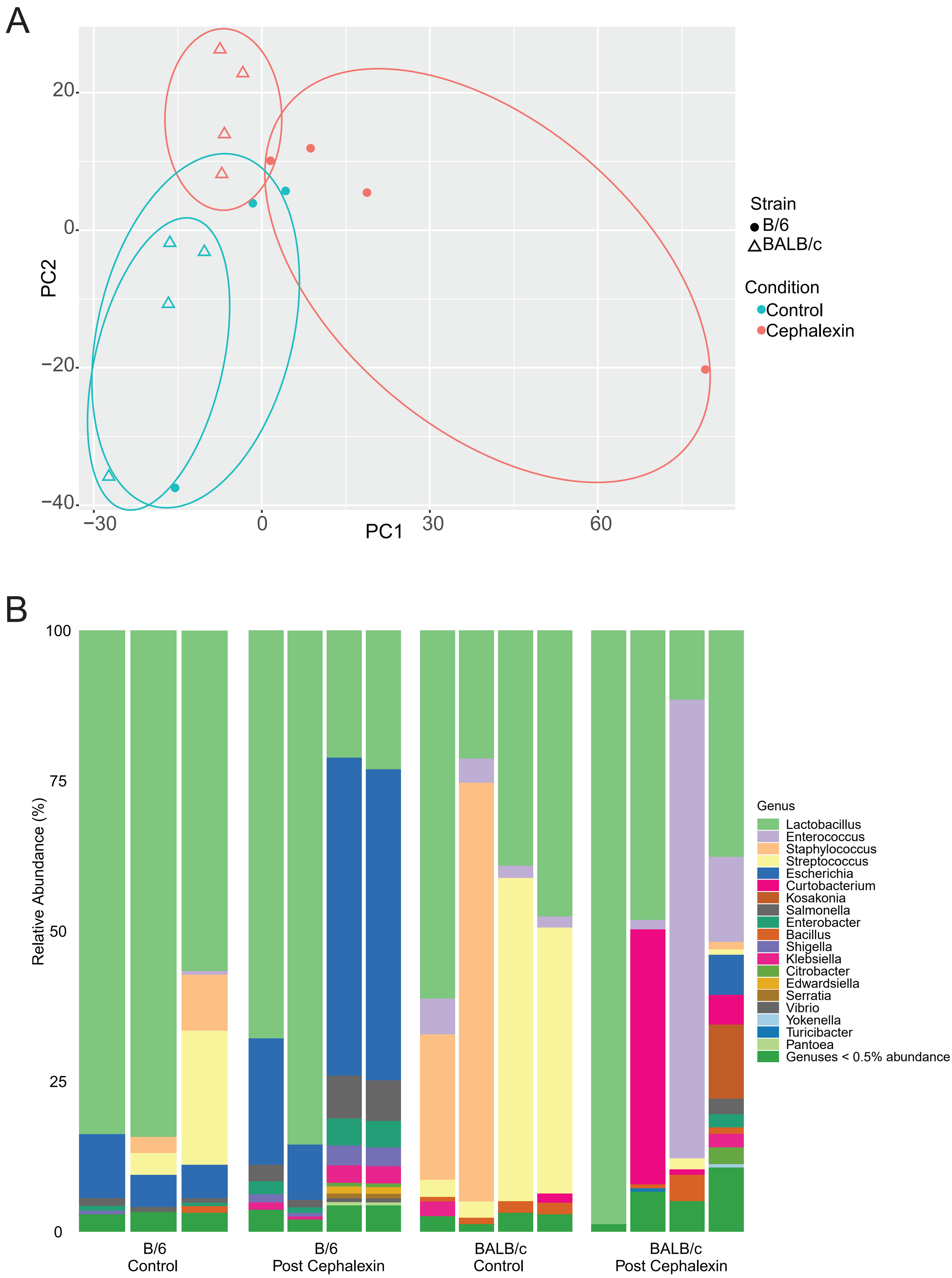
