## Supplementary Table 2 for "Disruption of maternal IgA by prenatal antibiotics precedes intestinal *E. coli* colonization and late-onset sepsis in neonates"

| | **Traditional Antibiotics** | **Non-Traditional Antibiotics** |
|---|---|---|
| Days antibiotics were given | 3.97 | 3.72 |
| Cephalosporin use | 0 | 20 |
| Gentamicin use | 0 | 19 |
| Vancomycin use | 0 | 4 |
| Clindamycin use | 0 | 21 |
| No azithromycin | 12 | 2 |

Supplemental Table 1: Neonatal outcomes following Maternal Antibiotics and PPROM,*Death, LOS, NEC, BPD/CLD, grade 4-3 IVH
